# Estimating Force of Infection from Serologic Surveys with Imperfect Tests

**DOI:** 10.1101/2020.06.09.20125724

**Authors:** Neal Alexander, Mabel Carabali, Jacqueline K Lim

**Affiliations:** MRC Tropical Epidemiology Group, London School of Hygiene and Tropical Medicine, London, United Kingdom; Department of Epidemiology, Biostatistics and Occupational Health, McGill University, Montreal, Quebec, Canada; Global Dengue and Aedes-transmitted Diseases Consortium (GDAC), International Vaccine Institute, Seoul, Korea

**Keywords:** Bayesian methods, diagnostic tests, force of infection, sensitivity, serology, specificity, surveys

## Abstract

The force of infection, or the rate at which susceptible individuals become infected, is an important public health measure for assessing the extent of outbreaks and the impact of control programs. Here we present methods for estimating force of infection from serological surveys of infections which produce lasting immunity, taking into account imperfections in the test used, and uncertainty in such imperfections. The methods cover both single serological surveys, in which age is a proxy for time at risk, and repeat surveys in the same people, in which the force of infection is estimated more directly. Fixed values can be used for the sensitivity and specificity of the tests, or existing methods for belief elicitation can be used to include uncertainty in these values. The latter may be applicable, for example, when the specificity of a test depends on co-circulating pathogens, which may not have been well characterized in the setting of interest. We illustrate the methods using data from two published serological studies of dengue.

---

The force of infection, or the rate at which susceptible individuals become infected, is an important public health measure used to assess the speed and extent of an epidemic, and the impact of disease control programs, as well as to prioritize and identify regions requiring further control, and vaccine implementation (1-5). For infections inducing lasting immunity, the force of infection is usually estimated via serological surveys (‘serosurveys’) of immunological status. Ideally, assays used in serosurveys should be highly sensitive and specific while also suitable for high throughput, in terms of the cost and personnel required (6-11). In practice, however, available assays may not completely meet all these criteria, as is currently evident with severe acute respiratory syndrome coronavirus 2 (SARS-CoV-2), the virus responsible for coronavirus disease (COVID19) (12).

The force of infection may be estimated from single or repeated serosurveys. In the former case, the simplest analysis is to assume that the force of infection was constant over calendar time and age, and consider age as the time at risk (13). More sophisticated models allow for changing force of infection over time, or over age, or even allow for maternal antibodies if the analysis includes new-borns or infants under a year of age (4, 14). Repeated surveys in the same individuals provide more robust estimates of the force of infection during a given study period (4, 7). Using repeated surveys, rate ratios can be obtained from binomial regression with complementary log-log link and the logarithm of the time between surveys as an offset (13). While age is used as the time at risk in the analysis of a single survey, in repeated surveys it can be considered a risk factor like any other. However, errors in test status are usually ignored, whether analysing one or more surveys. In particular, for repeat surveys, individuals testing positive at baseline are usually considered no longer at risk (1, 4, 7).

The choice of assay may substantially affect the study’s interpretation (15). Various methods have taken into account certain kinds of test imperfection, for either single or repeated surveys. In particular, Trotter & Gay (16) developed a compartmental model of multiple surveys, in which the force of infection and imperfect sensitivity were estimated for *Neisseria meningitidis*. For a single survey, Alleman et al. (17) and Hachiya et al. (18) estimated the force of infection, and simultaneously test sensitivity for rubella and measles, by assuming that imperfect sensitivity was the reason for seroprevalence not necessarily reaching 100% at the highest ages. Tan et al. (19) use a model for dengue, in which sensitivity reduces over time as antibody levels decrease, applied to two independent population serosurveys from blood donors. Olive et al. (20) estimated the force of infection for Rift Valley fever based on fixed values of sensitivity and specificity for a single survey. Here we provide methods to estimate force of infection, from a single serosurvey or two serosurveys in the same individuals, accounting for imperfect sensitivity and/or specificity, and uncertainty in these parameters.

## METHODS

We started from methods for estimating prevalence based on an imperfect diagnostic test, as reviewed by Lewis & Torgerson (21), and use similar notation. Estimation is done using a Bayesian framework and Markov chain Monte Carlo (MCMC) (22). We assume that the immune response being measured is long-lasting so that, for example, apparent seroreversions, i.e. changes over time from positive to negative, are due to test errors rather than loss of immunity. We use “seroprevalence” to mean the proportion of individuals with the underlying immune response, which the diagnostic tests measure with error.

### Model for single serosurvey

The probability of testing positive (*T* ^+^) is specified as a function of the unobserved true status (*π*), and the assumed values for sensitivity (*S*_*e*_) and specificity (*S*_*p*_):

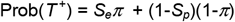

Then, representing a constant seroconversion rate, a binomial regression is specified with a complementary log-log link, and the logarithm of age as an offset. The only other term in the model is an intercept, which is the logarithm of the force of infection (13). A vague prior — Gaussian with mean zero and standard deviation 1,000 — is specified for the logarithm of the force of infection. Example data from a single serosurvey of dengue are from Colombo, Sri Lanka, which used a capture enzyme-linked immunosorbent assay (ELISA) to detect immunoglobulin G (IgG) (14, 23). Here we omit individuals aged less than six months to limit the influence of maternal antibodies.

### Model for two consecutive serosurveys in the same individuals

For two repeat serosurveys, priors are placed on the seroprevalences, and the values of interest are related via standard identities. The baseline seroprevalence is assigned a beta distribution with both parameters equal to 1, i.e. uniform on the interval [0,1]. The prior for the second seroprevalence is the same except that, consistent with the above assumptions, it is constrained to be at least as high as the baseline seroprevalence. For each survey, the positive and negative predictive values (*PPV* and *NPV*, respectively) are defined in terms of the seroprevalence and the assumed sensitivity and specificity:

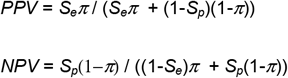

The probability of each person being truly seropositive, Prob(*D*^+^), is then *PPV* if the test is positive, and 1-*NPV* if the test is negative. The probabilities of testing positive or negative are functions of sensitivity and specificity, in the same way as for a single survey. Finally, the numerator of the force of infection is estimated as the increase in expected number of true positives from the first to the second survey, and the denominator is estimated as the expected person-time at risk, calculated as the sum of the individual times between the surveys, weighted by each individual’s probability of being seronegative at baseline. This is shown in the following equation, where the sum is over all individuals in both surveys, and the subscripts on *D* indicate the first or second survey:

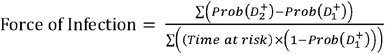

This is shown schematically, as a Directed Acyclic Graph, in Figure 1. Example data are from a community-based study of dengue in Medellin, Colombia, using a commercially available IgG indirect ELISA test (23). Residents were randomly selected, followed over time, and tested up to five times. For the current purpose, we use only the first survey, done in 2011, and the last one, done in 2014, approximately 26 months later.

**Figure 1.**
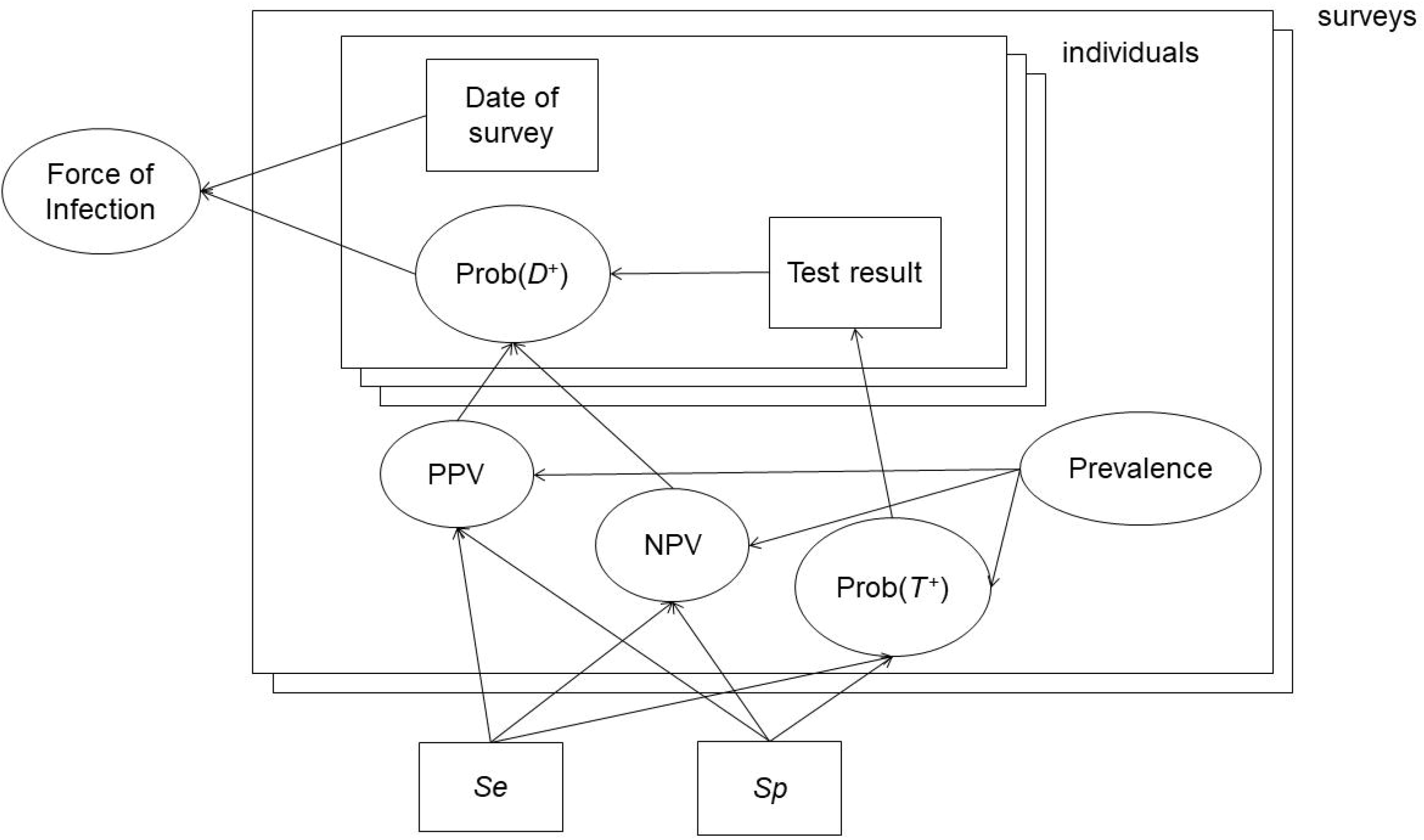
Directed Acyclic Graph (DAG) for the model for repeated serosurveys. The large rectangles show individuals nested within surveys. Both surveys and individuals have multiple stacked rectangles to show that there is more than one of each. The smaller rectangles represent data (results and times of tests) or model inputs (sensitivity and specificity). The other nodes are functions of the data, or of the unobserved seroprevalence, which is given a beta(1,1), i.e. uniform, prior. For individuals, Prob(*T* ^+^) indicates the probability of a positive test result and Prob(*D*^+^) indicates the probability of being truly seropositive.

In the standard binomial regression model for seroconversion across paired surveys, those individuals positive at baseline are assumed to be not at risk, i.e. there is no allowance for measurement error in the serostatus. By contrast, as well as seroconversion, the current model allows seroreversion, i.e. for individuals to change from seropositive to seronegative status.

### Uncertainty in sensitivity and specificity

Fixed values for sensitivity and specificity can be used for the repeat surveys, as for a single one. However, there may be reasonable doubt as to the exact values of sensitivity and specificity, e.g. because there cross-reacting pathogens circulate to an unknown extent. This uncertainty may have been quantified by systematic reviews, although their generalizability to a given setting may be doubtful. Another way to quantify uncertainty in terms of expert opinion, e.g. via the Delphi technique (24). Here we follow the elicitation method of Johnson et al. (25). For each parameter, each expert is presented with a range of values. For the current purpose, the parameters are sensitivity and specificity, each with a range of 0 to 100%, in intervals (“bins”) of 5%. Each expert is invited to i) make a point or “average” estimate of the parameter in question, then ii) indicate the upper and lower limit of their estimate, then iii) indicate their weight of belief by allocating a total of 100% over the bins, between the upper and lower limits, in units of 5%. So we have a total of six questions: three each for sensitivity and specificity. Johnson et al., used paper questionnaires and stickers for the units of 5% weight of belief. We adapted this to a spreadsheet in Microsoft Excel (Appendix 1). This approach could also be applied to the analysis of a single survey.

For the current study, beliefs were elicited from one of the authors (MC) who was also an investigator of the serological study in Medellin (23). In the case of dengue, one important consideration is whether the test in question may cross-react with other flaviviruses (26), or have lower specificity in those who have been vaccinated against them (27). The elicited distributions for sensitivity and specificity are used here to illustrate the current method and are not conclusive in terms of the performance of the test in question. Also, the considerations for other diagnostic tests and other settings will vary.

A smooth distribution between 0 and 1 was fitted to these belief weights. Both beta and logistic-normal families were fitted. Each has two parameters, which were fitted by the method of moments, i.e. equating the mean and variance of the belief weights to those of the distribution. The beta distribution was used for the estimation of the force of infection.

More broadly, some models for sensitivity and specificity are unidentifiable (28), i.e. not all the parameters can be estimated independently. For the current purpose, the estimated force of infection is evidently strongly associated with the sensitivity and specificity. Although posterior likelihoods of sensitivity, specificity and force of infection could be obtained from a formally consistent Bayesian model, the identifiability of such a model would need to be demonstrated. Our interest here is in information on sensitivity and specificity as inputs, not outputs. Hence, we have not referred to the elicited beliefs for sensitivity and specificity as “priors”. Although these beliefs are used in Monte Carlo simulation, posterior likelihoods are not obtained for them. Rather, values are repeatedly sampled from the fitted beta distributions of sensitivity and specificity, then MCMC estimation is done based on those values.

Credible intervals for a parameter are estimated as quantiles of samples drawn, via MCMC, from its Bayesian posterior distribution. Confidence intervals are quoted from frequentist analyses which were carried out for comparison.

### Software

We use the “rjags” package in R (version 3.6.3; The R Foundation for Statistical Computing). This package requires a separate installation of the JAGS package (29). R code is provided in Appendix 2. For the analysis of the Colombo survey, for each value of sensitivity and specificity used, a burn-in of 1,000 iterations was used, with estimates of the force of infection based on 50,000 iterations thinned by 10 (i.e. keeping every 10^th^ result). For the analysis of the repeat surveys in Medellin, the following was done separately for sensitivity and specificity: 2,000 draws were made from the fitted beta distribution then, for each draw, there was a burn-in of 2,000 and the force of infection was estimated from 5,000 iterations thinned by 50. This was done i) with sensitivity varying while holding specificity at 100%, ii) the reverse, and iii) with both parameters varying. Hence the distribution of the force of infection was estimated from 20,000 values. Assessment of convergence was done visually. For all MCMC models, the point estimate is taken to be the median of the iterative values and the 95% credible interval is from the 2.5^th^ to 97.5^th^ percentiles.

## RESULTS

### Single serosurvey

Figure 2 shows the fitted proportions of seropositive by age, in the dengue study in Colombo (14). Values of 85% sensitivity or specificity have been chosen to illustrate the method rather than on the basis of expert opinion or of comparison against a gold standard. However, they are in the range found for other dengue IgG ELISAs (30, 31). As expected, imperfect sensitivity implies higher seroprevalence, and imperfect specificity the reverse. The consequences of other values of sensitivity and specificity are shown in Figure 3. The confidence bands reflect sampling variability in the data rather than uncertainty in the values of sensitivity and specificity. From a standard frequentist analysis with binomial regression, the estimated force of infection is 13.7% per year (95% confidence interval 12.4-15.2%). Figure 3 shows that the results from the current model approach the results from the standard analysis as sensitivity and specificity tend to 100%. For 100% sensitivity the results from the current model are the same, and for 100% specificity the point estimate is the same and the credible interval is 0.1% lower (12.3-15.1%).

**Figure 2.**
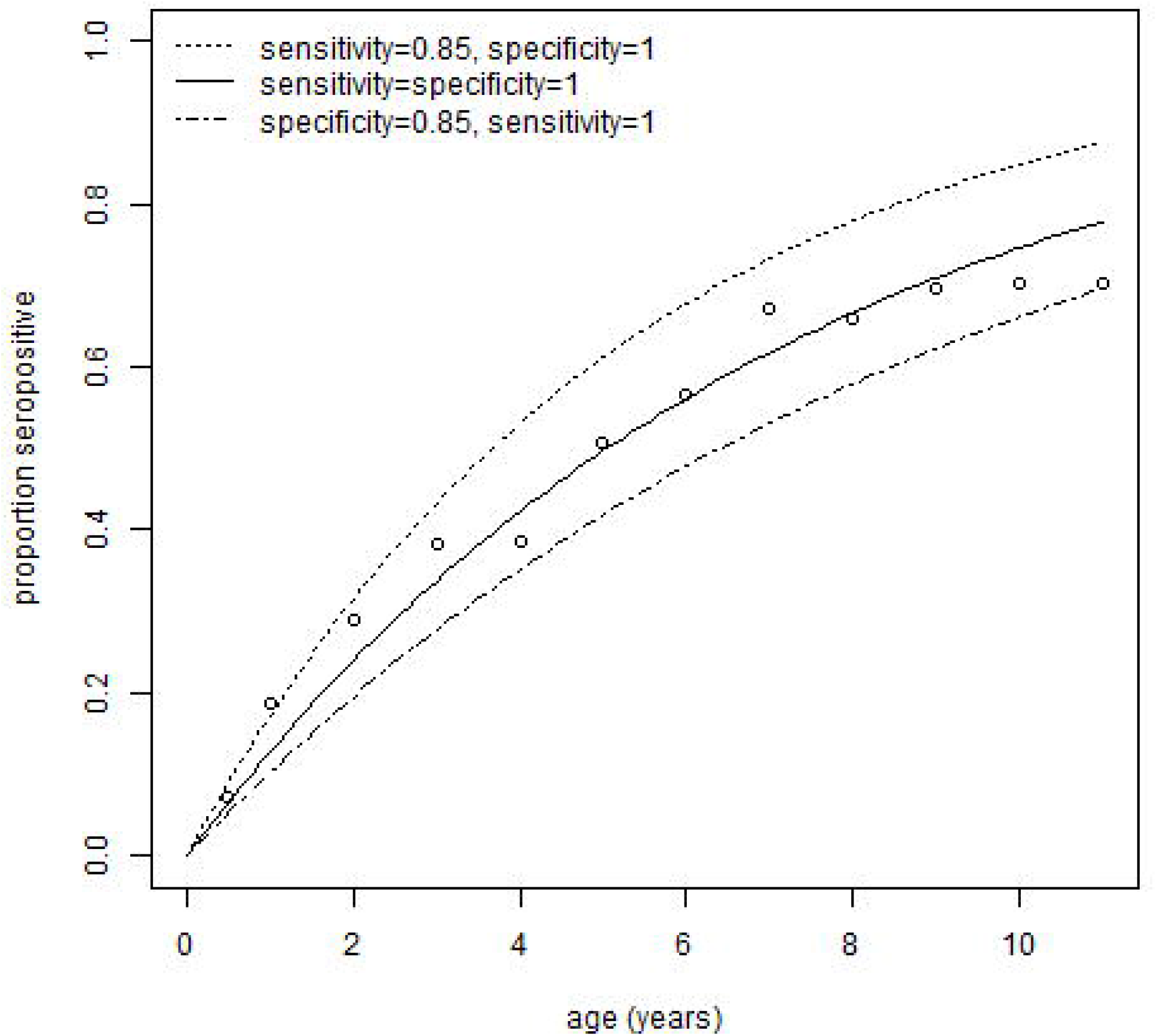
Proportion seropositive for dengue by age in Colombo (14). The solid line is the fit from a standard analysis assuming a perfectly sensitive and specific test. The upper dashed line is from an analysis assuming 85% sensitivity and 100% specificity, and the lower dashed line with these values exchanged.

**Figure 3.**
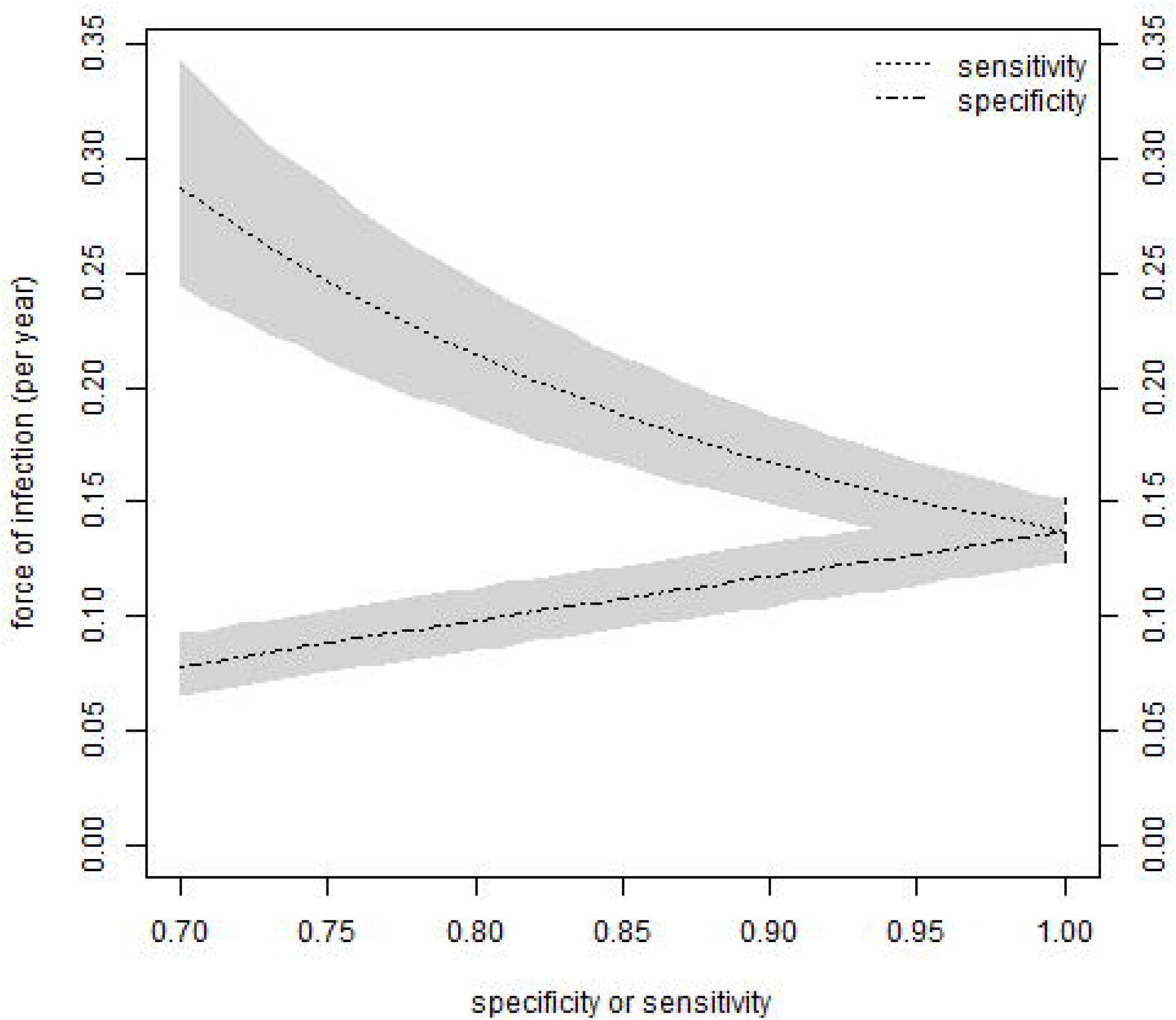
Relation between force of infection, sensitivity and specificity in the Colombo data. The force of infection is estimated for each value of sensitivity or specificity, considered fixed. In this figure, when sensitivity is less than 100% then specificity is assumed to be 100%, and conversely. The grey zones are the 95% credible intervals. As sensitivity and specificity approach 100%, to the right side of the plot, the credible intervals approach the 95% confidence interval from standard binomial regression (vertical dashed line).

### Two consecutive serosurveys

In Medellin, 705 people had test results available for both surveys (23). Of these, 260 originally tested negative, of whom 31 (11.9%) were positive on the second survey, approximately 26 months later. The remaining 445 originally tested positive, and all but four of these tested positive on the second survey. A standard frequentist binomial regression analysis, which was necessarily restricted to the 260 presumed at risk, estimates the force of infection as 5.9% per year with a 95% confidence interval from 4.0 to 8.2%.

Figure 4 shows the elicited distributions for the sensitivity and specificity of the dengue IgG ELISA used in the Medellin study. These are from only one expert and only used and intended for illustrative purposes. The distribution for specificity is closer to 100% and with lower variance than that for sensitivity. The figure also shows the fitted beta and logistic-normal distributions, the former of which was used to generate the force of infection results in Figure 5.

**Figure 4.**
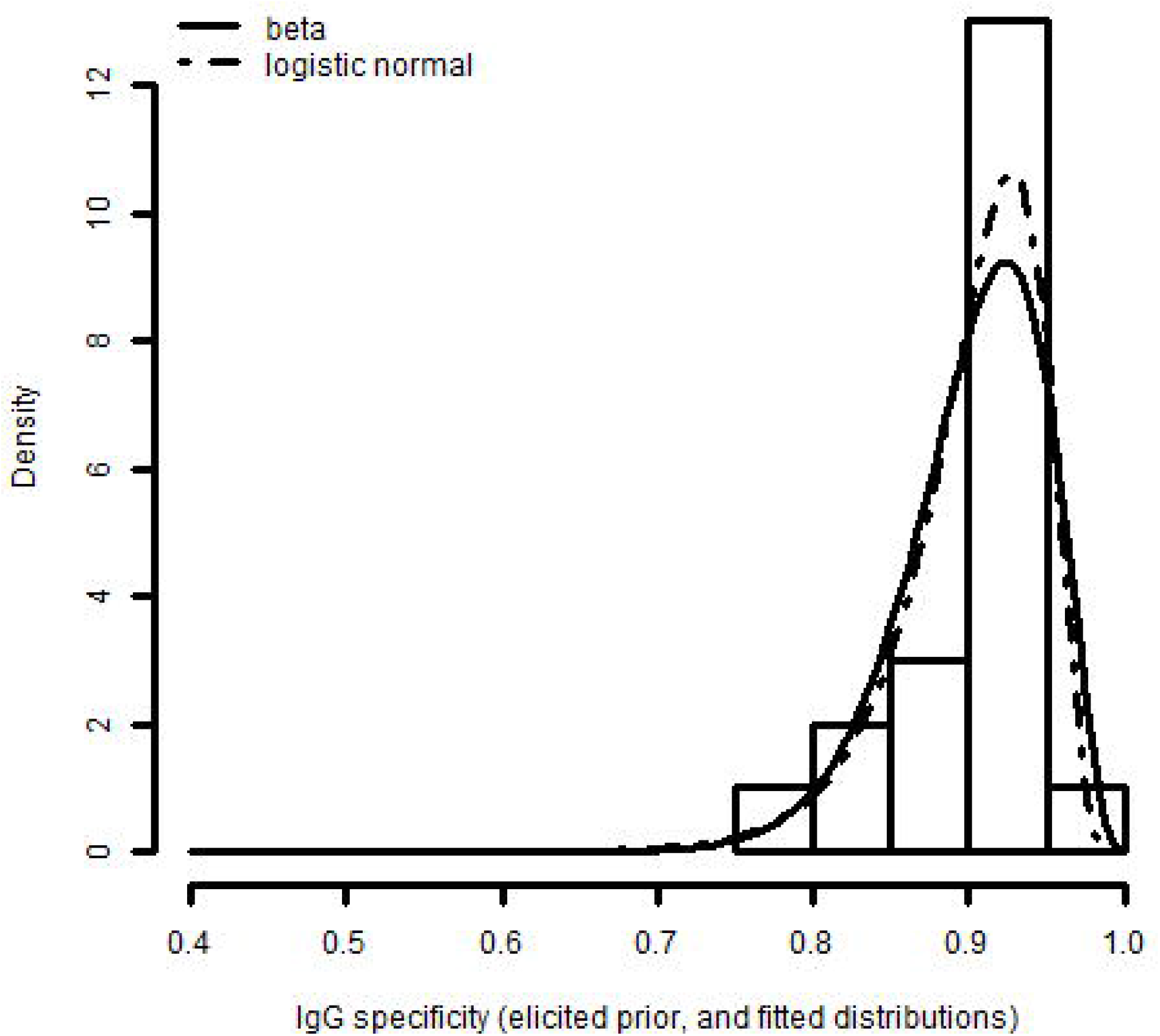
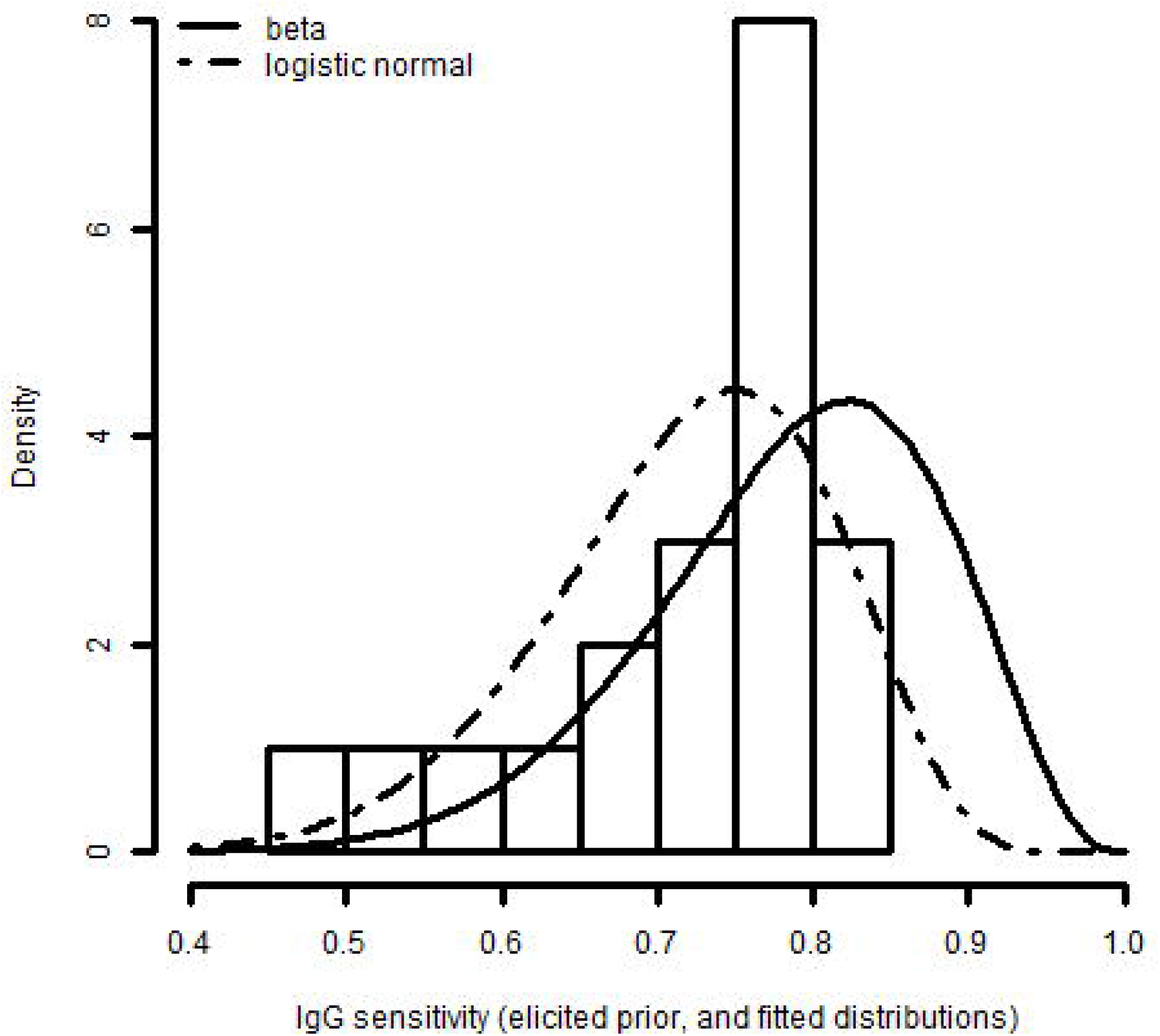
Uncertainty in a) specificity and b) sensitivity for Medellin study.

**Figure 5.**
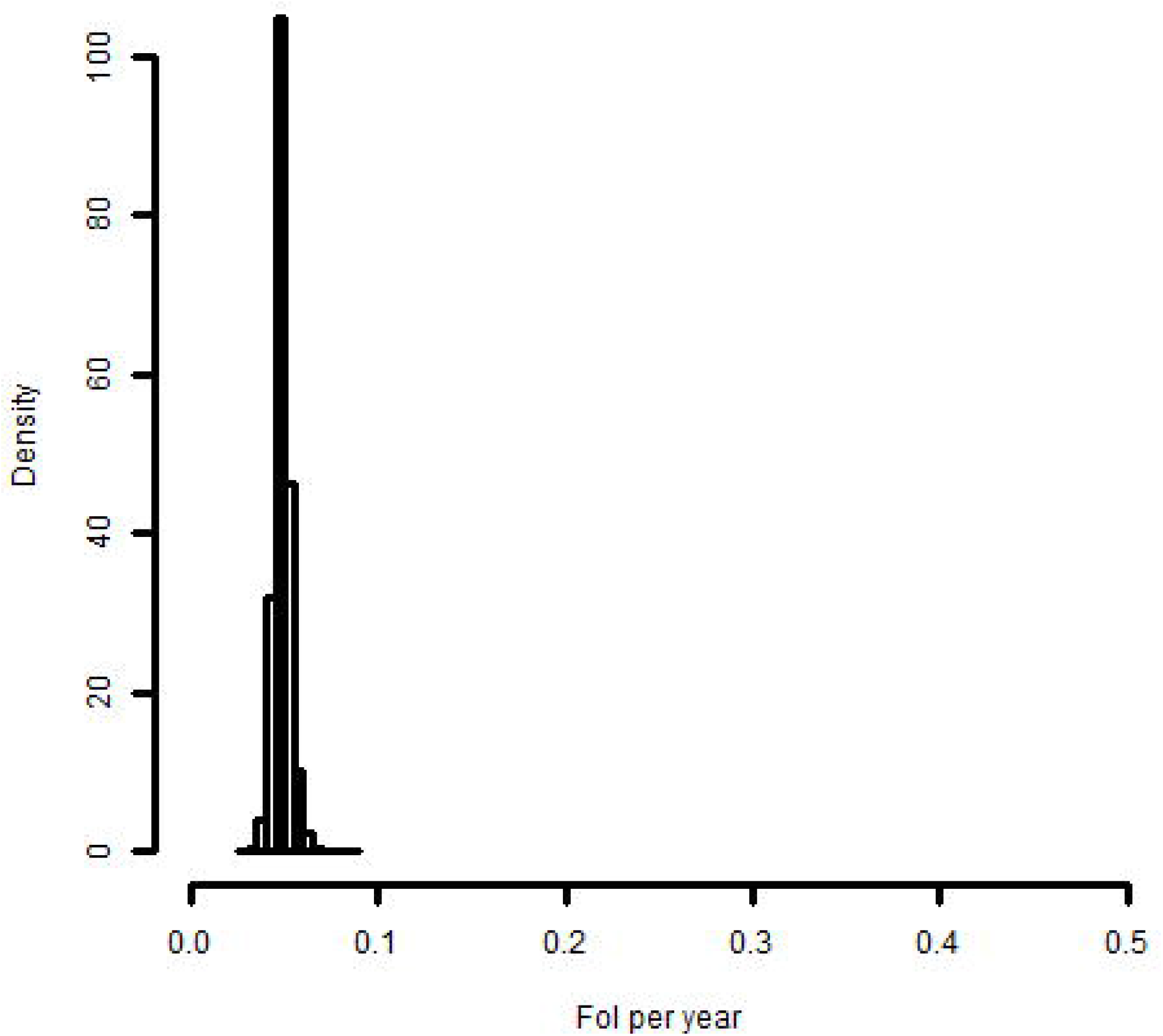
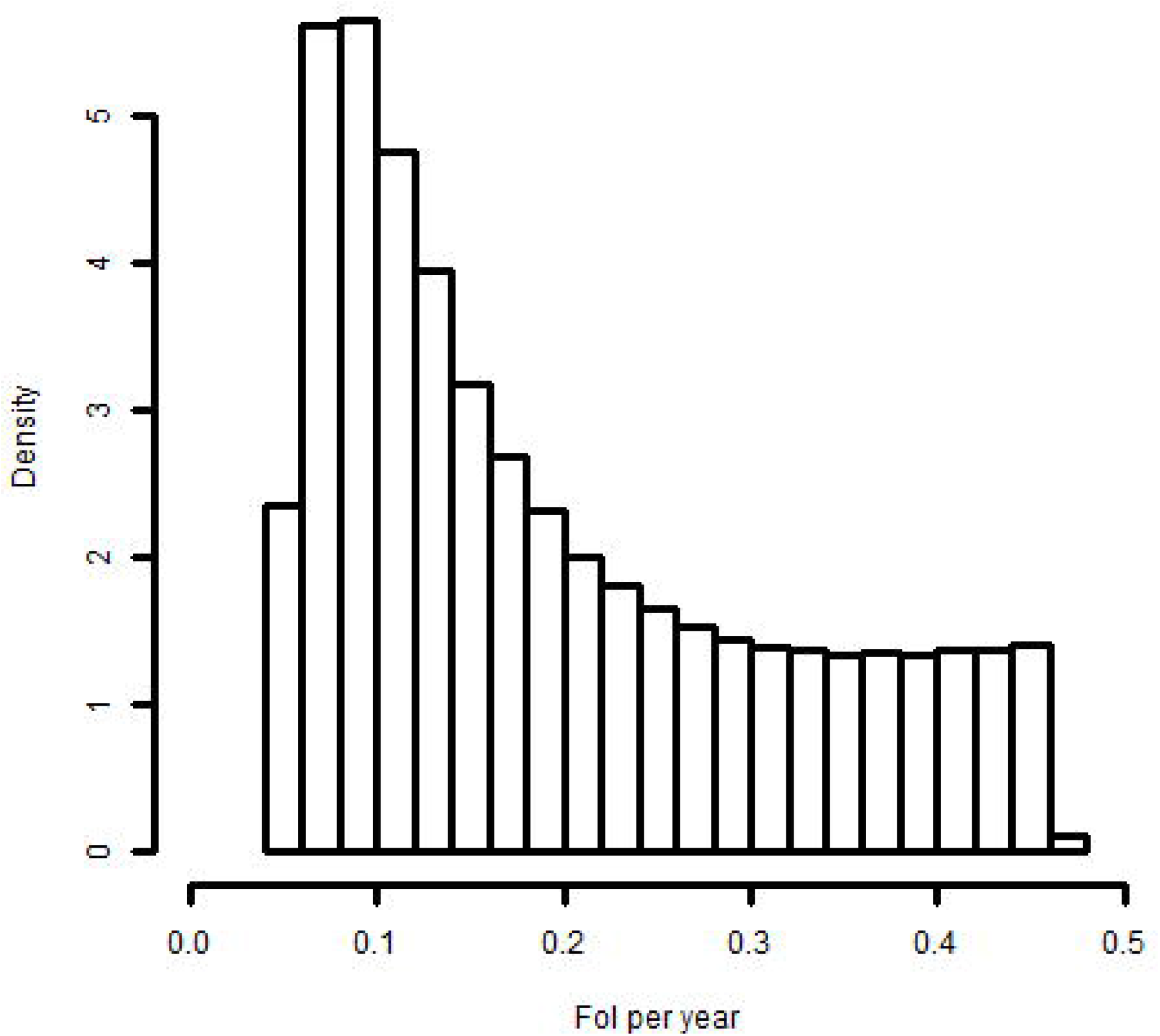
Posterior distributions of force of infection under a) varying specificity and b) varying sensitivity.

Results from the force of infection model are shown in Figure 5. As expected from Figure 4, the distribution of the force of infection taking into account uncertainty in specificity (Figure 5a) has smaller variation than that for sensitivity (Figure 5b). For the former, the point estimate of the force of infection is 4.8% per year, with a 95% credible interval from 4.0 to 5.8%. For varying sensitivity, the point estimate is much higher, at 15.6% per year, and the credible interval much wider: 5.5% to 44.4%. With sensitivity and specificity both varying, the results are qualitatively similar to Figure 5b (Appendix 3), the point estimate is again 15.6% per year, and the credible interval is from 4.9% to 44.5%.

## DISCUSSION

Not all assays are suitable for serological surveys. For example, the World Health Organization discourages the use of rapid tests in such studies of dengue (5), and the utility of serological assays for SARS-CoV-2 is currently being debated (12, 32). Statistical methods can help quantify the degree of uncertainty that would arise from the use of any given test. Previous studies have simultaneously estimated test sensitivity and force of infection for single or repeat surveys (16-18), and estimated the force of infection subject to fixed values for sensitivity and specificity in a single survey (20). Here we present methods for estimating the force of infection taking into account imperfect sensitivity and/or specificity, and uncertainty in these parameters, for either single or repeat surveys. Should well-established and generalizable values of sensitivity and specificity be available, they can be used in the methods described here. However, this is not always the case. For example, in the case of dengue, there may be cross-reaction with other flaviviruses (26), whose occurrence varies geographically.

The model for the single serosurvey, in which age is taken as the time at risk, applied to the dengue serosurvey in Colombo (14), showed how the force of infection depends on the assumed sensitivity and specificity. When perfect sensitivity and specificity are assumed, the results are effectively identical to those from the standard binomial regression. For the example of repeat serosurveys in Medellin (23), the elicited expert belief for the specificity was relatively precise, resulting in a fairly precise estimate of the force of infection (95% credible interval 4.0 to 5.8% per year). The belief for sensitivity was less precise and resulted in an interval estimate that was so wide (5.5 to 44.4%) as to potentially lack utility. The results from these two studies illustrate the method, but the force of infection values should not be taken as authoritative for the study settings.

We have opted for estimation in a Bayesian framework by MCMC (22). The model for a single serosurvey is similar to that of Lewis et al. (21) for prevalence, and may be soluble by direct application of maximum likelihood, hence avoiding the need for iterative sampling. The identifiability of some Bayesian models for the estimation of prevalence is affected by the choice of priors for sensitivity, specificity and other parameters: inaccurate priors can then give rise to inaccurate conclusions (28). Although it may be possible to ‘learn’ about both the assay parameters and the force of infection, here we have avoided identifiability concerns by including the elicited uncertainty in sensitivity and specificity via Monte Carlo simulation. In effect, the elicited distribution is both the prior and posterior distribution. This approach was shown for the model for repeat surveys but could equally be applied to the one for a single survey. It was illustrated by eliciting beliefs about sensitivity and specificity from a single expert. To reach substantive conclusions, multiple experts would be required (25). Estimates from systematic reviews could be used instead of expert opinion if they were generalizable to a given study area.

Future work could seek models with Bayesian priors for sensitivity and specificity, while still correctly estimating the force of infection. In the meantime, the use of Monte Carlo in an outer loop, with MCMC estimation each time, makes the analysis relatively time-consuming. Also, a reformulation would be required to allow the inclusion of covariates. The method is shown for two surveys, studies with more than two could be included, with each being constrained to have a seroprevalence no lower than the previous. Another limitation is the assumption that each individual has long-lasting immunity, so that apparent seroreversions are due to test errors rather than waning immunity. Depending on the infection in question, the validity of this assumption may depend on factors such as age and immunocompetence. In conclusion, the methods presented here can make more realistic estimates of force of infection, and can help inform the choice of serological tests for future serosurveys.

## Data Availability

The data that support the findings of this study are available from the corresponding author upon reasonable request.

## ACKNOWLEDGMENTS

Neal Alexander receives salary support from a) grant number MR/R010161/1 from the United Kingdom (UK) Medical Research Council and UK Department for International Development (DFID) under the MRC/DFID Concordat agreement, which is also part of the EDCTP2 programme supported by the European Union, and b) an award from FAPESP (2018/14389-0) and the UK Medical Research Council (MR/S0195/1) for the Brazil-UK Centre for Arbovirus Discovery, Diagnosis, Genomics and Epidemiology (CADDE). Mabel Carabali holds a Canadian Institutes of Health Research Banting Best Doctoral Award fellowship.

We are grateful to Oliver Brady, Philippe Mayaud and Paul Mee (London School of Hygiene & Tropical Medicine) for comments on an earlier version of this manuscript.

## Abbreviations

COVID-19: coronavirus disease 2019
DAG: directed acyclic graph
ELISA: enzyme-linked immunosorbent assay
IgG: immunoglobulin G
MCMC: Markov chain Monte Carlo
*NPV*: negative predictive value
*PPV*: positive predictive value
*S*_*e*_: sensitivity
SARS-CoV-2: severe acute respiratory syndrome coronavirus 2
*S*_*p*_: specificity

## FIGURE LEGENDS

**Appendix 1**

Excel file for use in eliciting beliefs.

**Appendix 2**

R code. One file for single survey, one for repeat surveys, and one with utility functions.

**Appendix 3**

Figure: estimation of the force of infection in the Medellin study, with uncertainty in both sensitivity and specificity.

